# Inactivated Poliovirus Vaccine Induces Antibodies that Inhibit RNA Synthesis of SARS-CoV-2: An open-label, pre-post vaccine clinical trial

**DOI:** 10.1101/2021.10.05.21264598

**Authors:** Brittany A. Comunale, Erin Jackson-Ward, Yong Jiang, Laura P. Ward, Qianna Liu, Lyuqing Ji, Michelle Lai, Lilly Engineer, Roderick A. Comunale, Qiyi Xie

## Abstract

**Background:** Poliovirus vaccination induces an adaptive humoral immune response; *in vitro* experiments show polio-immune sera contain antibodies against the poliovirus RNA transcriptase that cross-react with SARS-CoV-2. While structural similarities between poliovirus and SARS-CoV-2 could have major implications for the COVID-19 response worldwide, polio-induced immune responses against SARS-CoV-2 have not been confirmed in prospective clinical trials.

**Objective:** To evaluate whether immune sera from adults who recently received inactivated poliovirus vaccination (IPV) can block SARS-CoV-2’s ability to synthesize RNA.

**Intervention:** IPV intramuscular injection.

**Measurements:** Pre-inoculation and 4-weeks post-inoculation sera were tested for anti-3D^*pol*^ (RNA-dependent RNA polymerase, RdRp) antibodies using enzyme-linked immunosorbent assays (ELISA). To assess IPV’s ability to induce antibodies that inhibit SARS-CoV-2 RNA synthesis, immune-based detection assays tested RdRp enzymatic activity in polio-immune sera.

**Results:** 298 of the 300 enrolled participants completed both on-site visits. Comparing pre-inoculation to 4-week samples, 85.2% of participants demonstrated an increase in anti-3D^*pol*^antibodies against RdRp proteins. Among tested post-inoculation samples, 94.4% demonstrated inhibition of SARS-CoV-2 RNA synthesis. Few inoculation-related side effects were reported (2.0%), all were minor.

**Limitations:** Participants were not systematically tested for COVID-19, though known exposures were reported and positive results (1.7%) were documented.

**Conclusion:** IPV can induce antibodies that inhibit SARS-CoV-2 RNA synthesis, minimizing the risk of viral replication in infected individuals. This finding has practical implications for resource-deficient areas that may have limited access to newly developed COVID-19 vaccines and/or areas with low COVID-19 vaccination rates due to hesitancy.

**Funding Source:** Private donors.

**Registration:** ClinicalTrials.gov: NCT04639375.

## Introduction

SARS-CoV-2, the virus that causes COVID-19, has affected 221 countries and territories with over 235 million cases and 4.8 million deaths as of October 3, 2021 **[1]**. Considering the prevalence, infectivity, and virulent characteristics of COVID-19, rapid identification of the viral agent allowed unparalleled efforts to produce COVID-19-specific vaccines **[2,3]**. However, despite emergency use authorization of four vaccines (Pfizer-BioNTech, Moderna, Johnson & Johnson, and AstraZeneca), all shown in trials to be more than 71% effective at preventing hospitalization due to COVID-19 **[4,5]**, only 34% of the world has been fully vaccinated, allowing the persistence of viral spread and creation of threatening variants **[6]**. While the reasons for suboptimal vaccination rates are multi-factorial, in developed countries, they include hesitancy regarding the efficacy and safety due to perceptions of unusually rapid development and approval; in other settings, the issues include inability to access vaccines and/or lack of infrastructure to support administration (further complicated by the need for refrigeration and/or two doses for some options). It would likely be of great public interest if an existing vaccine with established safety, production, and administration procedures on a global scale were to be able to provide anti-COVID-19 activity.

The poliovirus vaccine may offer such an option. During vaccine development efforts exploring SARS-CoV-2 specific targets, the possibility that existing vaccines for viruses with similar molecular structures and/or mechanisms to SARS-CoV-2 might induce antibodies against COVID-19 infection was raised **[7-9]**. Exploratory and serological data specifically support poliovirus inoculation as a potential prophylactic measure against SARS-CoV-2 infection **[10,11]**. The poliovirus vaccine contains a non-structural protein, nsp-12, or RNA-dependent RNA polymerase (RdRp), which is responsible for directing RNA synthesis and viral replication in RNA viruses, including SARS-CoV-2 **[12-14]**. RdRp is closely related to the N-protein, a key structural protein necessary for SARS-CoV-2 viral assembly and replication that has been targeted for SARS-CoV-2 diagnosis **[15-17]** and potential therapeutics **[18,19]**. As such, RdRp has been at the forefront of developing antiviral drugs, such as Remdesivir, which has been used to treat patients hospitalized with COVID-19 **[20,21]**. Due to its ability to induce antibodies that respond to RdRp antigens, the poliovirus vaccine is coming to the fore as a possible tool for COVID-19 protection.

A recent retrospective serological study, prompted by observed structural commonalities between RdRp proteins in poliovirus and SARS-CoV-2 **[22]**, further supports the theory that poliovirus vaccination may be capable of creating an immunological response against COVID-19. Initial findings demonstrate that SARS-CoV-2 RdRp antibodies have the capacity to recognize one or more epitopes of the poliovirus RdRp protein, 3D^*pol*^. Thus, the poliovirus vaccine, which contains the 3D^*pol*^ (RdRp) protein, induces antibodies that may cross-react with SARS-CoV-2. Preliminary data suggest that inactivated poliovirus vaccination (IPV) could induce heterologous immunity, indicating that memory T cells could identify shared epitopes between SARS-CoV-2 and poliovirus. An effective immune response to the invading SARS-CoV-2 antigen may then be elicited and theoretically disrupt the viral replication process. The study results indicate proteins other than “spike” proteins (i.e., RdRp) may be appropriate targets for immunity and vaccine development, but additional clinical data would be beneficial for further validation **[11]**.

While observational studies and *in vitro* experiments of polio-immune sera have been examined retrospectively in the context of the COVID-19 pandemic, the immunological association between poliovirus vaccination and SARS-CoV-2 has not been evaluated prospectively. The present study discusses the outcomes of an open-label, pre-post clinical trial that evaluated inhibition of SARS-CoV-2 RNA synthesis by polio-immune sera in adults.

## Methods

### Trial Design and IRB Approval

A single arm, open-label, pre-post vaccine clinical trial (ClinicalTrials.gov: NCT04639375) was conducted between November 2020 and June 2021 in San Diego, California, in accordance with International Conference on Harmonisation Good Clinical Practice (ICH GCP) and the United States (US) Code of Federal Regulations (CFR) applicable to clinical studies (45 CFR Part 46, 21 CFR Part 50, 21 CFR Part 56, 21 CFR Part 312, and/or 21 CFR Part 812). IRB approval (Advarra IRB) was obtained for the study protocol, informed consent forms, recruitment materials, and all subject-facing materials. All participants provided written informed consent. The trial was conducted in compliance with ICH GCP and the ethical standards described in the WMA Declaration of Helsinki.

### Participant Enrollment

Three hundred volunteers between the ages of 18 and 80 years with no active infectious disease, no previous history of COVID-19, and no prior COVID-19 vaccination (Emergency Use Authorized vaccines available at the time of enrollment) were entered into the study. Participants were recruited through internet advertisements, referrals, and clinicaltrials.gov.

Participants of child-bearing potential were required to have a negative pregnancy test prior to being vaccinated and to be willing to use an effective method of birth control from the time of entry into the study to 30 days following vaccination. Exclusion criteria included: known allergic reactions to components of the poliovirus vaccine, febrile illness within 14 days, positive test result for SARS-CoV-2 antigenemia at any time prior to screening, positive test result for SARS-CoV-2 antibodies at any time prior to screening, treatment with an investigational drug or other intervention within 90 days prior to enrollment in the study, inoculation with poliovirus vaccine within the last 12 years, any symptoms related to SARS-CoV-2 infection (fever or chills, cough, shortness of breath, fatigue, muscle or body aches, headache, new loss of taste or smell, sore throat, congestion, runny rose, nausea, vomiting, diarrhea) within 48 hours, and women who are pregnant or breast-feeding.

### Intervention and Follow-Up

At the first on-site visit (Day 1), enrolled participants provided a baseline blood sample (preinoculation) and subsequently received one dose (0.5 mL) of inactivated poliovirus vaccine (IPV; IPOL, Sanofi Pasteur) by intramuscular injection. Blood specimens collected pre-inoculation were tested for baseline titers of poliovirus antibodies including RdRp. Between Day 3 and 7 post-inoculation, study staff contacted participants via phone to document any potential side effects or adverse events from the recent immunization. Participants also kept a daily temperature log to record any instances of fever for one week following vaccination. At the second on-site visit (28 ± 3 days post-inoculation), additional blood specimens were collected and tested for IPV-induced poliovirus antibodies including RdRp, hereafter called “anti-3D^*pol*^ (RdRp) antibodies,” as well as potential inhibition of SARS-CoV-2 RNA synthesis via impeding RdRp enzymatic activity. While on-site visits for this study have been completed, COVID-19 status will continue to be monitored for all enrolled participants at both six and twelve months post-inoculation (through May 2022).

### Outcomes

The primary objective was to evaluate whether IPV could induce polio antibodies in adults. The secondary objective was to evaluate whether polio-immune sera could inhibit SARS-CoV-2 RdRp functionality, or inhibit the virus’ ability to synthesize RNA and promote viral replication.

### Immune Profiling

For the primary outcome, commercially available enzyme-linked immunosorbent assay (ELISA) kits were used to measure anti-3D^*pol*^ (RdRp) antibody titers for both baseline (Day 1) and postinoculation (Day 28) human sera. Standard laboratory procedures were followed to perform the ELISA tests. In brief, IPV with 3D^*pol*^ (RdRp antigen) was coated on the microplate wells, followed by an application of the human serum sample to the plate. Antigen-specific antibodies in the serum sample then bound to the wells, and unbound antibodies were washed out with PBS-T buffer. Next, a solution of horseradish perodixase (HRP) conjugated goat anti-human IgG/A/M secondary antibody was added against the human antibodies to each of the wells. Unbound enzyme conjugated antibodies were then washed out with PBS-T buffer. TMB, a chromogenic substrate, was then added to each well. Once the TMB substrate turned blue by the HRP enzyme on the secondary antibody, 2M of sulfuric acid were added to the wells, changing the solution color to yellow, ultimately ceasing the reaction. An ELISA plate reader quantified the reaction activity by reading O.D 450nm absorbance. The relative serum response units, referred to as “arbitrary units,” were calculated using a nonlinear regression analysis and a 4-parameter fitting algorithm from the ‘drc’ package provided by R statistical software **[23]**.

### Measuring Inhibition of RNA Synthesis in Polio-Immune Sera

For the secondary outcome, SARS-CoV-2 RdRp functionality or RNA synthesis was measured *in vitro* from post-inoculation (Day 28) human sera by an adapted method that was originally created for Zika virus RdRp **[24]**. A streptavidin-biotin system was used to detect the enzymatic activity of the SARS-CoV-2 RdRp enzyme, nsp-12, which is a catalyst for RNA synthesis in SARS-CoV-2 **[22,25]**. The ELISA plates were coated with streptavidin to capture biotinylated oligodT complementary binding with de novo synthesized poly(A) RNA fragments, which were catalyzed by SARS-CoV-2 RdRp enzymes that were purified from insect cells.

Fifty-four of the 298 post-inoculation serum samples were randomly selected to be tested with the RdRp polymerase assay, due to the expensive nature of immune-based detection methods. Proportionality and two-tailed t-tests were used to compare characteristics of the randomly-tested subset to those of the overall cohort.

The assay plate setup included one negative control (in which all components including the standard serum diluted 1:16 were present, except for the RdRp enzyme), one positive assay control, and one positive sample control (complete RdRp enzymatic reaction system, but no serum present). The positive sample control’s enzymatic activity (O.D 450 nm) was set at 100% as the reference comparison; each experimental sample was individually compared to the sample control, and the serum sample’s inhibition effects of RdRp were calculated as a percentage of the positive sample

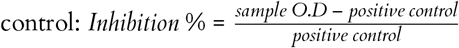

The recorded intensity corresponded with the SARS-CoV-2 RdRp functionality, or the virus’ ability to synthesize RNA, which would enable normal viral replication. A positive value (inhibition %) demonstrated up-regulated RdRp activity (signifying RNA synthesis progressed normally, which would lead to an infected individual getting sick) and a negative value reflected downregulated activity (indicating RNA synthesis was inhibited, thereby disrupting viral replication and minimizing an exposed individual’s risk of experiencing viral effects). An ELISA plate reader was used to record the activity and raw data were analyzed on Microsoft Excel.

## Results

A total of 298 of the 300 enrolled participants (age *m* = 51.1 years; 53.7% female) completed both of the on-site visits in San Diego, California **(Figure 1; Table 1)**. One participant died before the second on-site visit, though the cause of death was determined to be unrelated to COVID-19 and to the study by both the PI and medical monitor; a second participant was lost to follow-up.

**Table 1.** Characteristics of Study Participants.

| Characteristic | Overall Cohort (n = 300) | Randomly-Tested Subset (n = 54) | P Value |
| --- | --- | --- | --- |
| Age – yr |  |  | 0.11 |
| Mean | 51.1 | 54.9 |  |
| Median | 54.0 | 60.0 |  |
| Range | 18-80 | 18-80 |  |
| Sex – no. (%) |  |  |  |
| Male | 139 (46.3) | 24 (44.4) | 0.80 |
| Female | 161 (53.7) | 30 (55.6) | 0.80 |
| Race/Ethnicity – no. (%) |  |  |  |
| Caucasian | 181 (60.3) | 31 (57.4) | 0.69 |
| Hispanic/Latino | 75 (25.0) | 16 (29.6) | 0.47 |
| Asian | 28 (9.3) | 5 (9.3) | 0.98 |
| Black/African American | 12 (4.0) | 2 (3.7) | 0.92 |
| Native Hawaiian/ Pacific Islander | 2 (0.7) | 0 (0.0) | 0.55 |
| American Indian/ Alaska Native | 2 (0.7) | 0 (0.0) | 0.55 |
| Other | 2 (0.7) | 0 (0.0) | 0.55 |

\*The P values for sex and race/ethnicity were based on proportionality tests; those for age were based on two-tailed t-tests.

**Figure 1.**
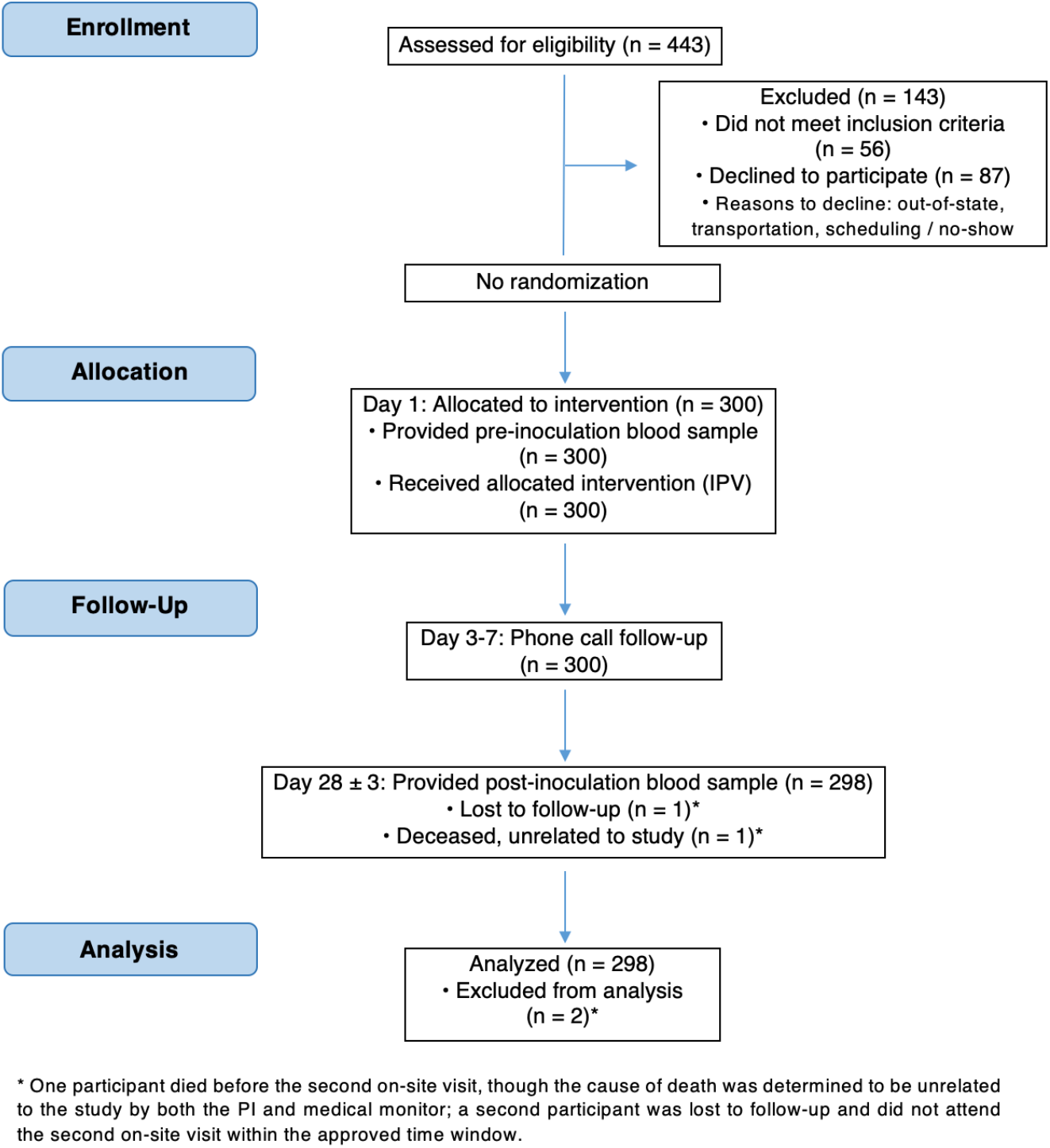
Study Design. Three hundred participants were enrolled in the single arm, open-label interventional study. All 300 participants received IPV, provided baseline blood specimens, and participated in the Day 3-7 follow-up phone call. In total, 298 participants provided post-inoculation blood specimens, which were then analyzed for preversus post-inoculation titer comparisons of anti-3D^*pol*^ (RdRp) antibodies.

### Reported Side-Effects

The Day 3-7 phone call follow-up included an assessment of side effects or adverse events from the immunization. Across all 300 calls, five participants (1.7%) reported experiencing mild swelling for about 24 hours near the site of injection. At the second on-site visit (Day 28 ± 3), participants returned the daily temperature logs they used to record any instances of fever they may have experienced during the first week following vaccination. One participant (0.3%) reported a low-grade fever (100.7°F), which lasted about 24 hours. No other adverse events or side effects related to immunization were reported.

### Self-Reported COVID-19 Status

Five participants (1.7%) reported testing positive for COVID-19 before the second on-site visit (Day 28 ± 3). In all five cases, participants were not experiencing serious symptoms themselves, but were aware of contact with someone who had tested positive for COVID-19, and independently pursued testing. One participant remained asymptomatic; the other four reported mild symptoms (i.e., fever, fatigue, congestion, sore throat), which only lasted 1-3 days. COVID-19 status will continue to be monitored for all enrolled participants through May 2022. As of August 25, 2021, no participant in this trial (0.0%) has been hospitalized or died due to COVID-19.

### IPV-Induced Immunity: Antibody titers pre- and post-inoculation

Overall, there was a significant increase in levels of anti-3D^*pol*^ (RdRp) antibodies, comparing baseline sera (Day 1), referred to as “pre-inoculation,” to sera collected 28 ± 3 days post-inoculation (two-tailed t-test, p<0.001; **Figure 2**). Immune responses varied across participants, as 85.2% of the paired serum samples demonstrated an increase in antibody titers, 12.4% showed a decrease, and 2.3% had no change **(Table 2)**. The participants who did not demonstrate an increase in levels of anti-3D^*pol*^ (RdRp) antibodies had naturally high antibodies pre-inoculation (significantly higher than baseline titers in the naturally low antibody group, two-tailed t-test, p<0.001). Conversely, participants who had naturally low titers before immunization reflected a significant increase in antibody levels post-immunization (two-tailed t-test, p<0.001). The demographics of the seven participants (2.3%) who did not exhibit an immune response to the inoculation were as follows: age, *mean* (range) = *52* (25-70) years; 57.0% male; 42.8% Caucasian, 28.6% African American, 14.3% Hispanic, 14.3% Asian.

**Table 2.** Titers of Anti-3D^*pol*^ (RdRp) Antibodies.

| Titer Change | No. of Samples (%) | Pre-Inoculation Mean (SD) | Post-Inoculation Mean (SD) | Mean % Change (SD) | P Value |
| --- | --- | --- | --- | --- | --- |
| Increase | 254 (85.2) | 1.07 (0.7) | 2.09 (2.3) | 122.04 (192.3) | 1.70E-13 |
| No Change | 7 (2.3) | 1.05 (0.8) | 1.05 (0.8) | 0.00 (0.0) | 1.00 |
| Decrease | 37 (12.4) | 2.07 (1.4) | 1.64 (1.5) | 17.08 (26.2) | 1.08E-4 |

\*P values were based on two-tailed t-tests.

**Figure 2.**
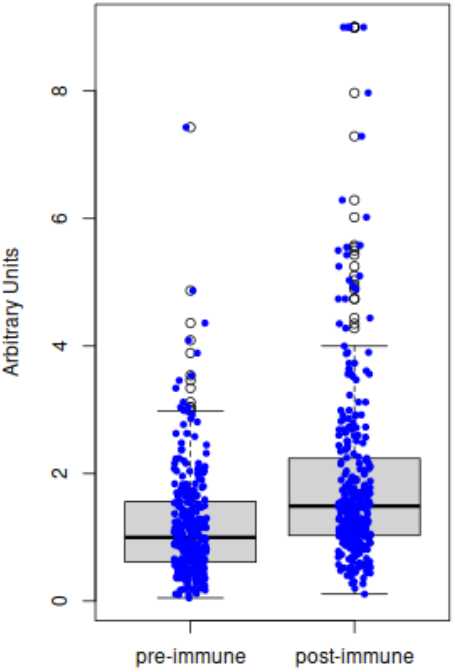
Comparison of antibody titers pre-inoculation (Day 1) versus post-inoculation (Day 28±3). The results from ELISA of the 298 paired serum samples demonstrate that inoculation with inactivated poliovirus vaccine (IPV) significantly increases the titers of anti-3D^*pol*^ (RNA-dependent RNA polymerase, RdRp) antibodies in adults (two-tailed t-test, p<0.001).

### Polio-Immune Sera Inhibit RNA Synthesis of SARS-CoV-2 via Impeding RdRp Enzymatic Activity

Fifty-four of the 298 post-inoculation serum samples were randomly selected to be tested with the SARS-CoV-2 RdRp polymerase assay. As shown in Table 1, there were no significant differences in demographics between the overall cohort (n=300) and randomly-tested subset (n=54). Among the 54 samples, 51 (94.4%) exhibited negative values or down-regulated RdRp activity, indicating SARS-CoV-2’s RdRp functionality, or ability to synthesize RNA, was inhibited. While there was a variable extent of inhibition across individuals, the recorded trend of negative values indicates IPV has the capability to minimize the risk of disease progression in infected individuals. Three of the tested samples (5.5%) exhibited positive values or up-regulated RdRp activity, signifying normal RNA synthesis, which would lead to viral replication and cause an infected individual to become sick. These results show polio-immune sera are very likely to inhibit SARS-CoV-2 RNA synthesis to some extent, thereby affecting the viral replication process.

## Discussion

Based on findings from this prospective study, inactivated poliovirus vaccination increases the level of anti-3D^*pol*^ (RdRp) antibodies in adults, especially among those with low baseline titers, and most notably, polio-immune sera are capable of inhibiting RNA synthesis of SARS-CoV-2.

IPV can induce polio antibodies in adults. While antibody titers increased for 85.2% of participants, 12.4% demonstrated a decrease in anti-3D^*pol*^ (RdRp) antibodies. Such occurrences are expected, though, as these participants already had a very high level of antibodies at baseline, which were comparable to the level of antibodies observed in the post-inoculation group that demonstrated an increase in titers. When individuals have a high level of pre-existing antibodies, they are less likely to develop a robust immune response post-inoculation **[26,27]**.

Polio-immune sera can inhibit SARS-CoV-2 RdRp functionality, or inhibit the virus’ ability to synthesize RNA and promote viral replication. Approximately 95% of tested polio-immune sera inhibited SARS-CoV-2 RNA synthesis. Since viral replication requires RNA synthesis, IPV’s demonstrated ability to inhibit this process in SARS-CoV-2 has large clinical implications for the COVID-19 pandemic.

These findings further validate the results from a recent retrospective serological study, in which sera from adults vaccinated with IPV showed positive trends of mounting antibodies against polio RdRp that cross-reacted with SARS-CoV-2 RdRp **[11]**. These statistically significant immune responses have the potential for clinically significant impact, as IPV-induced antibodies have demonstrated the ability to inhibit SARS-CoV-2 RNA synthesis, indicating that IPV may minimize the risk of viral replication or disease progression in individuals infected with SARS-CoV-2. This preliminary evidence suggests that if confirmed in a randomized controlled vaccine trial for preventing COVID-19 infection, hospitalization, and death, IPV may be a viable supplement to existing COVID-19 vaccines. This readily-available vaccine could be an invaluable means for protection in resource-deficient countries that may not have access to other COVID-19 vaccines. The extent of such protection may vary across individuals, but further investigations are currently underway regarding the differentiation in protection both among and between sub-populations, as well as the range of protection as it relates to SARS-CoV-2 variants.

These study results may be generalized to populations similar to the studied cohort—predominantly Caucasian or Hispanic/Latino, middle-aged adult individuals. Although 90% of the world’s population has already received the poliovirus vaccine as part of childhood vaccination programs, the present results are only applicable to people who have been immunized with IPV within the last 28 days. Since antibodies wane over time, adults who were once immunized as children with the poliovirus vaccine, as seen in this study, may no longer have antibodies in the present day **[28,29]**.

Due to the inclusion criterion specifying an absence of “active infectious disease,” the studied cohort can be considered more representative of the general population than those in comparative COVID-19 vaccine studies, which only allow for participants to be of optimal health. In this study, participants with underlying medical conditions, such as diabetes, were still considered eligible to participate, enhancing generalizability of the presented serological data. There was an opportunity to include a broader population for this study, as IPV is an FDA-approved vaccine with an existing safety profile, differentiating it from newly developed vaccines that have to investigate safety and efficacy in their respective trials.

Since this clinical trial was the first study of its kind to evaluate IPV in the context of the COVID-19 pandemic, the study design was intentionally simple as a single arm, open-label study, and thus includes inherent limitations that should also be considered opportunities for future studies. As a single-site study, the sample size, geographic representation, and demographics were limited. While the studied cohort as a whole was representative of the local San Diego population, Asian and Hispanic/Latino subgroups were slightly underrepresented. In response, future studies may conduct a larger trial, both in terms of size and through a multi-centered approach, in order to enhance generalizability across different populations. There are already active follow-up serological studies focused on stratifying immunological responses across different ages and racial/ethnic groups, but future studies may expand the current understanding of varying immune responses in more diverse settings.

In addition to expanding in size, future trials may also incorporate more complex design features to build upon this preliminary study. A multi-armed approach could be used to compare populations of varying vaccination statuses, such as COVID-19 vaccine only, IPV only, COVID-19 vaccine and IPV, and placebo. In this study, enrolled participants had not received any prior COVID-19 immunizations, though if they wanted to get vaccinated, they could do so after their second on-site visit. Future studies should also implement routine COVID-19 testing, measure antibody titers beyond the 28-day period to see when antibodies may wane (i.e., 6 or 12 months), and test immune responses and RNA synthesis inhibition across different IPV dosing regimens, as the recommended dosage for adults traveling to high-risk polio regions can vary from one to three doses **[30]**.

As the poliovirus vaccine has been used across the globe for over six decades, both for childhood immunizations and for travel protection, it has a well-established history and extensive safety profile. The prospective serological data presented here suggest IPV may be considered for another purpose, to provide some protection against COVID-19. While larger studies, including randomized vaccination trials evaluating COVID-19 outcomes, should further validate this claim, the preliminary data from this study reinforce findings from a prior retrospective study **[11]**. Additionally, this study encourages stronger consideration of using IPV as a potential therapeutic agent that is more readily available in resource-deficient countries, as well as a well-established vaccine that may be viewed as a safer and more widely trusted option for vaccine-hesitant populations. With the ever-evolving COVID-19 pandemic and imminent, malicious variants, any opportunity to further boost the population’s health should be greatly considered.

## Data Availability

The original contributions generated for this study are included in the article/supplementary material, further inquiries can be directed to the corresponding author.

## Acknowledgements

We would like to acknowledge and thank Robin J. Larson, MD, MPH of the Geisel School of Medicine at Dartmouth for providing mentorship and commentary on the manuscript, Anafrancesca Comunale, MSc, MBA, Esq. for leading administrative operations, and the clinical research team of RAC, Inc. for conducting the presented study.

